# Mental health and weight loss in men: an exploratory mixed methods study of the Game of Stones trial

**DOI:** 10.1101/2024.12.12.24318850

**Authors:** Claire E Torrens, Katrina Turner, James Swingler, Catriona O’Dolan, Alice MacLean, Lisa Macaulay, Stephan U Dombrowski, Alison Avenell, Seonaidh Cotton, Michelle C McKinley, Kate Hunt, Cindy Gray, Frank Kee, Graeme MacLennan, Pat Hoddinott

## Abstract

**Objective:** Weight management interventions can affect mental health. Mental health can affect engagement with weight loss interventions or services. This study explored mental health and wellbeing outcomes, retention and participant experiences of mental health within the Game of Stones trial.

**Methods:** Mixed methods process evaluation within a 3-group randomised controlled trial: behavioural text messages with financial incentives, texts alone, and waiting list control, for 585 men with obesity. Secondary outcomes analysed descriptively included: Warwick-Edinburgh Mental Wellbeing Scale, Weight Self-Stigma Questionnaire, EQ-5D-5L, EQ-5D-5L anxiety and depression subscale, Patient Health Questionniare-4, and retention. Three categories of participants were compared: ever diagnosed with a mental health condition (n=146; 25.0%), latent mental health condition (n=142; 24.3%) no mental health condition (n=295; 50.6%). Semi-structured interviews (n=54) were conducted after 12 months and analysed using Framework method.

**Results:** A higher proportion of men who self-reported ever having a mental health condition had a disability, multiple long-term conditions, were under financial strain and were single compared to those with those with a latent mental health condition and no mental health condition. Improvements from baseline were shown for weight stigma, wellbeing and PHQ-4 at 12 months for men in intervention groups with a mental health condition and latent mental health condition. EQ-5D-5L Visual Analogue Scale scores improved across all mental health categories and trial groups, but EQ-5D-5L and EQ-5D-5L-AD scores were inconsistent. Retention at 12 months was 76.0% (mental health condition), 70.4% (latent mental health condition) and 72.5% (no mental health condition). The qualitative evidence indicated that stress, anxiety and depression were experienced in different ways by men during the programme. Mental health difficulties were unique to the individual, could be episodic, recurrent, cyclical or ongoing and were a barrier to behaviour change for some but not for others.

**Conclusion:** The trial was able to engage and retain men regardless of mental health category. Behavioural text messages with or without incentives helped some men lose weight, but not others. Observed heterogeneity for mental health and wellbeing measures is problematic for weight management trials with men.

## Introduction

The World Health Organisation estimates that worldwide around 890 million people are living with obesity(1) and around 1 in 8 people live with a mental health condition(2). The determinants of and inter-relationships between obesity, mental health and wellbeing and health inequalities are complex. Obesity is associated with a higher prevalence of mental health conditions, including depression and anxiety(3–5). Furthermore, people with mental health conditions have an increased risk for obesity, diabetes and cardiovascular disease compared to the general population(6). Health inequalities are apparent for people with mental health conditions and people living with obesity, with the Covid-19 pandemic further highlighting disparities(7).

Weight management interventions can improve both physical and mental health. A systematic review of adult weight management interventions found greater improvements in depression, mental health related quality of life and self-efficacy in intervention groups compared to control groups(8). However, the impact of offering a financial incentive to lose weight on mental health has received little attention in recent systematic reviews(9, 10). Fewer men than women engage with weight loss services(11) and men with mental health conditions have lower engagement with services than men without(12). In addition, men tend to under-report mental health conditions compared to women in all age categories(13) and use different terms when describing mental health and wellbeing(14). It is recommended that weight loss interventions are specifically designed and tailored for men(12).

Game of Stones was a randomised clinical trial of a digital weight management intervention designed with men, for men(15). The trial recruited 585 men, aged 18 or over with a Body Mass Index (BMI) equal to or greater than 30kg/m^2^, via General Practice, community and social media strategies between July 2021 and May 2022 from centres in and around three UK cities. Participants were randomized to three groups: daily behavioural text messages with financial incentives; text messages alone and a waiting list control group. The texts included evidence and theory-based behaviour change techniques based on the Health Action Process Approach(16), self-determination theory(17), and the behaviour change maintenance model(18). Texts were not designed to improve mental health, however mental health and wellbeing was included within some weekly themes. Examples of the text messages are available(15).

The financial incentive design drew on the theory of behavioural economics showing that people ascribe more value to something when it belongs to them (endowment effect) and are more motivated to avoid losses than they are to achieve similarly sized gains (loss aversion)(19, 20). Men randomised to the text messages with financial incentives group were informed £400 had been placed in an account for them, and if they met all three weight loss targets (i.e. 5% at 3 months, 10% at 6 months, maintaining 10% at 12 months) they would receive the full £400 after their 12 month follow up appointment. If a target was not met, they would lose part of this £400.

The two primary outcomes for the trial were: within participant weight change between baseline and 12 months, comparing each intervention group with the control group. Men in the texts with incentives group had significantly improved weight loss compared to the control group (mean difference, −3.2%; 97.5% CI, −4.6% to −1.9%; P < .001). The weight loss in the text messages alone group was not significantly greater than the control group (mean difference, −1.4%; 97.5% CI, −2.9% to 0.0, P = .05).

There is no consensus for which mental health and wellbeing measures to use for men participating in weight management trials in the community. Public and Patient Involvement (PPI) in the trial told us that mental health and stigma are key issues for people desiring weight loss. For questionnaires men preferred a wellbeing rather than a “mental health condition” focus and advised consideration of the negative burden of longer depression/anxiety measures(20). The wellbeing measures selected with PPI were Warwick-Edinburgh Mental Wellbeing Scale (WEMWBS)(21, 22) and the Weight Self-Stigma Questionnaire (WWSQ)(23). The anxiety and depression measures selected were Patient Health Questionnaire-4 item (PHQ-4)(24) and the Anxiety and Depression subscale of the Euro-QOL 5 Dimension five-level (EQ-5D-5L-AD)(25). NICE Guidance CG123(26) recommends using the PHQ-4 questionnaire in community settings to ascertain whether a more detailed mental health assessment is required(24).

Analysis showed that there were no statistically significant differences in WEMWBS, WWSQ, EQ-5D-5L, EQ-5D-5L-AD or PHQ-4 scores between the text messages with financial incentives group compared to control; or between text messages alone and control at 12 months(15). Text messages with financial incentives significantly improved the EQ-5D Visual Analogue Scale by 5.00 points (97.5% CI, 0.76, 9.25, P = .008), compared to control; text messages alone improved the EQ-5D Visual Analogue Scale by 3.71 points (97.5% CI, -0.75, 8.16, P = 06) compared to control, which was not significant. Of the 585 men in the trial, 146 (25%) participants self-reported a mental health condition at baseline, 227 (39%) participants lived in the two more deprived postal code quintiles for the Index for Multiple Deprivation(15). Subgroup analysis showed that measures of disadvantage, mental health or wellbeing status at baseline did not moderate the primary outcome of percentage weight lost between baseline and 12 months(27).

To inform future trials and further understand the relationship between text messaging, with or without financial incentives, and mental health, this paper aims to 1) explore trial retention by mental health category 2) compare different mental health and wellbeing measures at baseline and after the 12 month intervention by mental health category and 3) understand how men living with mental health conditions experienced the Game of Stones trial.

## Methods

Mental health and wellbeing data for WEMWBS, WSSQ, EQ-5D-5L, and PHQ-4 were collected at baseline and 12 months as part of a self-report participant questionnaire available in paper and electronic formats. The PHQ-4 was added as a new secondary outcome after the trial start, commencing in November 2021, but before collection of 12-month follow-up data(28).

Three categories of mental health status were generated from baseline data and applied to both quantitative and qualitative data:

1. *Mental health condition (MHC)*. All participants were asked ‘Has your doctor <u>ever</u> told you that you have/had a mental health condition?’ to which they needed to answer yes.
2. *Latent mental health condition (LMHC)*. This category aimed to identify participants that may have undetected mental health conditions or be at risk of developing one in future. This was defined as men who did not self-report a previous/current doctor diagnosed mental health condition but had a high score on at least one of the following measures: PHQ-4 (score 3+); EQ-5D-5L-AD (score 4+); WSSQ (score 42+); or a low score on WEMWBS (score 14-40). The cut-off scores for the PHQ-4, WSSQ and WEMWBS respective measures are guided and derived from the work of: Löwe (2010)(24); Lillis (2010)(23); and Bianco (2012)(22) respectively. The EQ-5D-5L-AD levels are level 1 not anxious or depressed; level 2 slightly, level 3 moderately, level 4 severely and level 5 extremely anxious or depressed. Levels 4 and 5 were decided as the appropriate cut off by the research team.
3. *No mental health condition (No MHC/LMHC)*. This category consists of participants who answered “no” to the question ‘Has your doctor ever told you that you have/had a mental health condition?’ and had scores that were not in the *latent mental health condition* range. We refer to these categories as mental health condition, latent mental health condition and no mental health condition hereafter.

A mixed methods process evaluation for men living with multiple long-term conditions and/or mental or physical disability was undertaken in parallel to this paper, which details methods for how long-term conditions and disability were identified(29). UK country specific measures for the Index of Multiple Deprivations were used, which describe participant postcode area of residence according to five deprivation quintiles(15).

### Quantitative methods and analysis

All exploratory quantitative results are presented as frequency and percentages for categorical variables, and as the mean, standard deviation and count for numerical variables. The observed differences in results between the three categories of mental health status are descriptive, with no formal statistical analyses undertaken.

### Qualitative methods and analysis

Semi-structured interviews were held with men in the two intervention groups following their 12-month assessment. These explored men’s views and experiences of the intervention components (i.e. texts, weight loss targets and financial incentives) and of being in the trial. A topic guide (S5 Topic Guide). was used to ensure consistency across interviews. This included questions on the perceived impact of health and wellbeing on participation, engagement and weight loss. Men in the text and incentives group were asked additional questions about their experience of being offered money to meet weight loss targets.

The interviews were audio-recorded and transcribed verbatim. Men who had consented in writing to being approached for interview were purposively sampled: by intervention group, trial centre and whether or not they were living with a mental health condition or multiple long-term conditions.

Three members of the research team (CT, CO and KM), all of whom had qualitative research experience, interviewed men they had not met during the trial. The interviews were held remotely by telephone or online via Zoom or Microsoft Teams to encourage participation and reduce participant burden. Verbal consent was recorded at the time of interview.

The qualitative analysis was guided by the five stages of Framework method(30): familiarisation; identifying a thematic framework; indexing; charting; mapping and interpretation. The analysis was theory driven, expanding on findings from the previous feasibility study(19, 20, 31). All steps of the analysis were carried out by the same 3 researchers (CT, CO and AM), with support from the process evaluation group (KT, KH, PH, SD, CMG) at each stage of the process. An initial thematic framework was developed by researchers after reading a diverse sample of early interviews and identifying key themes. This framework was then developed further with contributions from members of the trial’s process evaluation group and subsequently tested with additional transcripts. Once the thematic framework had been agreed, all transcripts were uploaded to NVivo v20 and divided among researchers for the process of indexing, charting and mapping. Matrices were used to summarise and chart data by themes and sub-themes. Attributes were assigned for intervention group, deprivation and health and wellbeing (e.g. mental health condition) as well as key outcome data (e.g. weight loss trajectory: met one or more targets or met no targets). The matrices helped the researchers to identify patterns with the aim of contextualising and interpreting participant outcomes. Data collection and initial analysis proceeded in parallel, and data collection ended when the data had adequate ‘power’ to address the study objectives(32).

### Retention, harms and benefits data

Reasons for non-attendance at appointments or withdrawing from the trial were documented by researchers where they were known. Researchers also recorded descriptive data on how weight measurements (e.g. in person on study scales, by video or by questionnaire using own scale) were collected on the trial case report forms. Harms and benefits data were collected for all trial participants at weight assessments: 3 months, 6 months and 12 months for intervention groups; and at 12 months only for the control group. Researchers asked: “Has anything unexpected happened since your last appointment as a result of taking part in the study (these can be either helpful or harmful consequences)?”. Methods for eliciting harms and unexpected benefits, and results by trial group are reported elsewhere(15). These data were analysed in excel spreadsheets, which were read and re-read by PH, KT and LM to identify themes and patterns within the data. Harms and benefits data were triangulated and reported with qualitative data as a final analysis step to further inform understanding of participant experiences.

Ethical Approval for the Game of Stones trial was received from the North of Scotland Research Ethics Committee 2 [20/NS/0141].

## Results

Baseline data for mental health were available for 583 of the 585 participants: 146 (25.0%) participants reported having ever had a mental health condition, 142 (24.4%) had a latent mental health condition and 295 (50.6%) had neither, falling into the no mental health condition category. Key baseline characteristics are presented in Table 1 with the full baseline characteristics in S1 table.

**Table 1.**
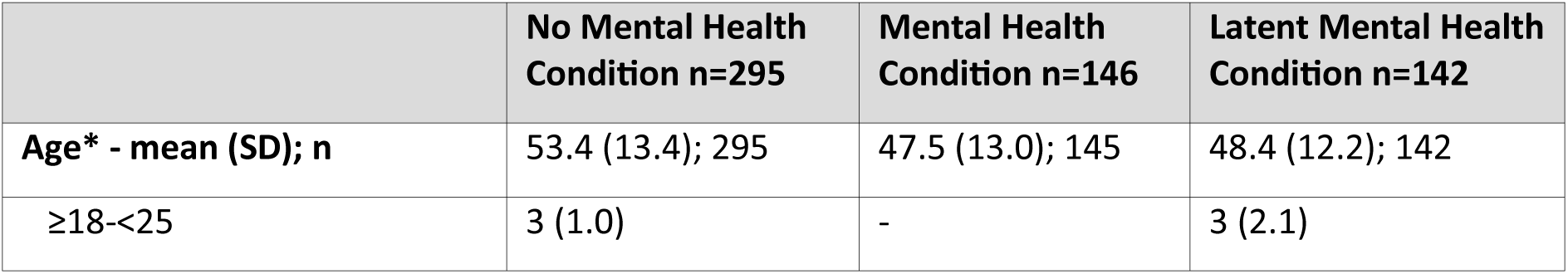
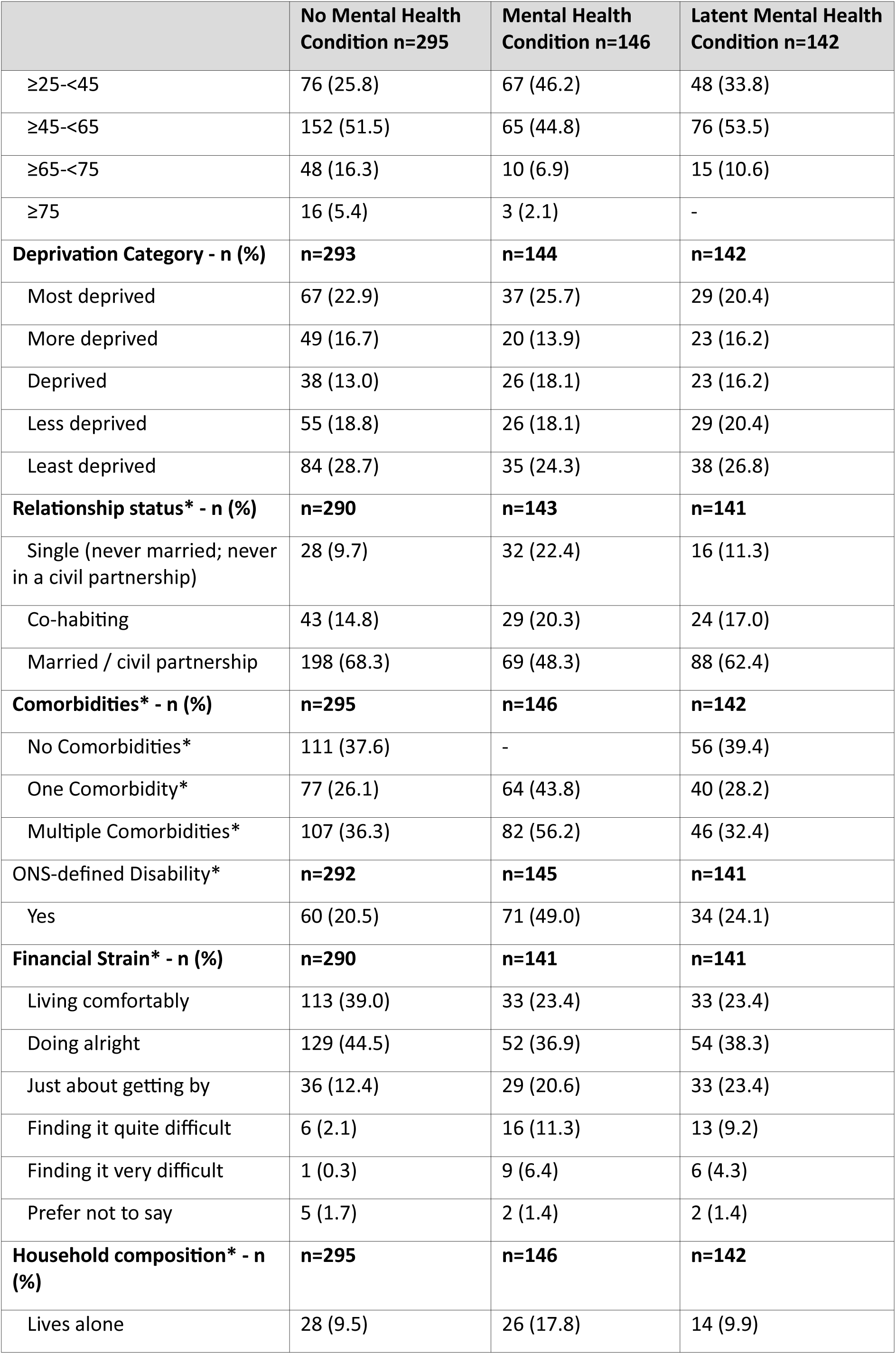
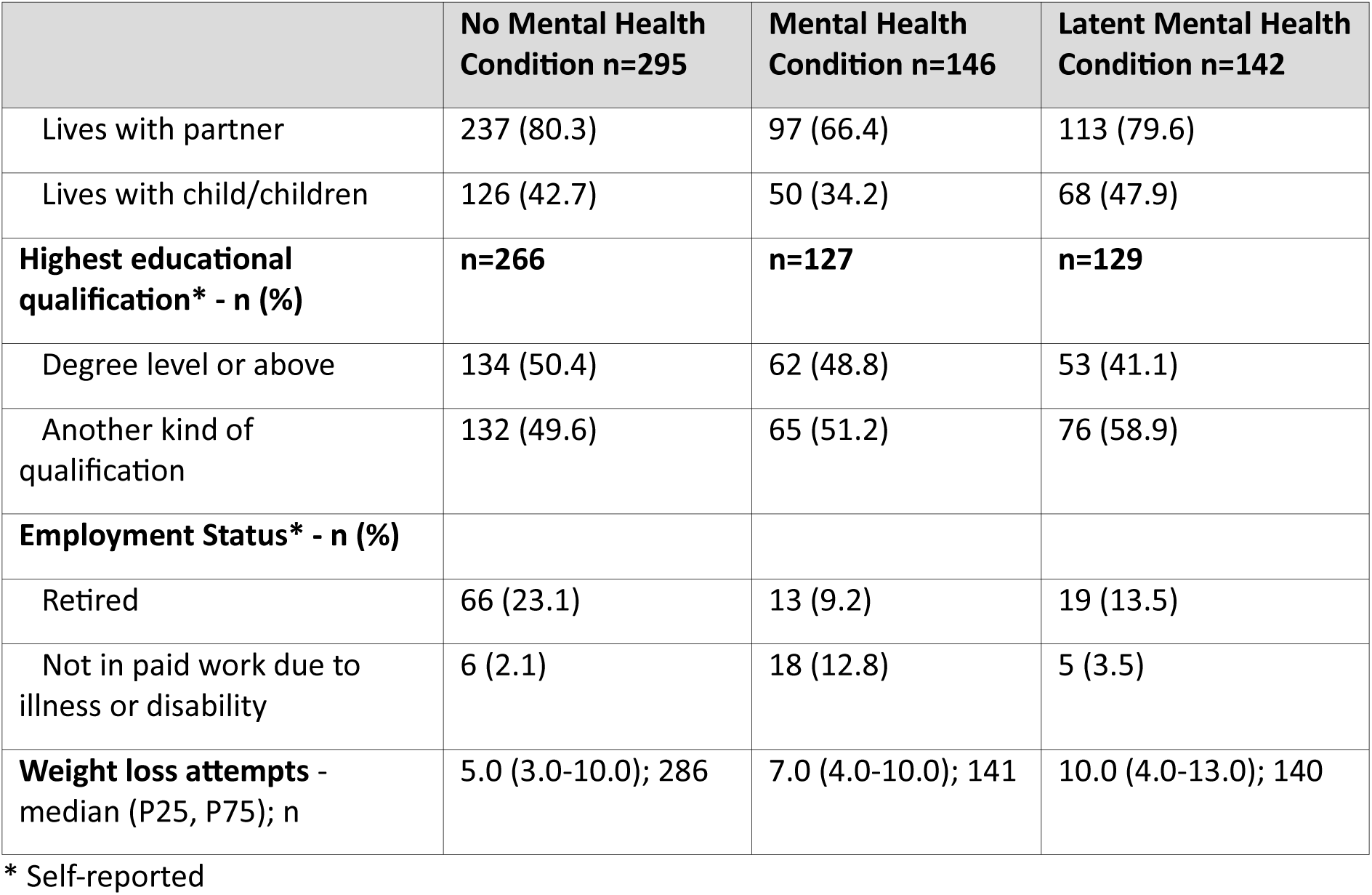
Baseline characteristics according to mental health category.

**Table 2.**
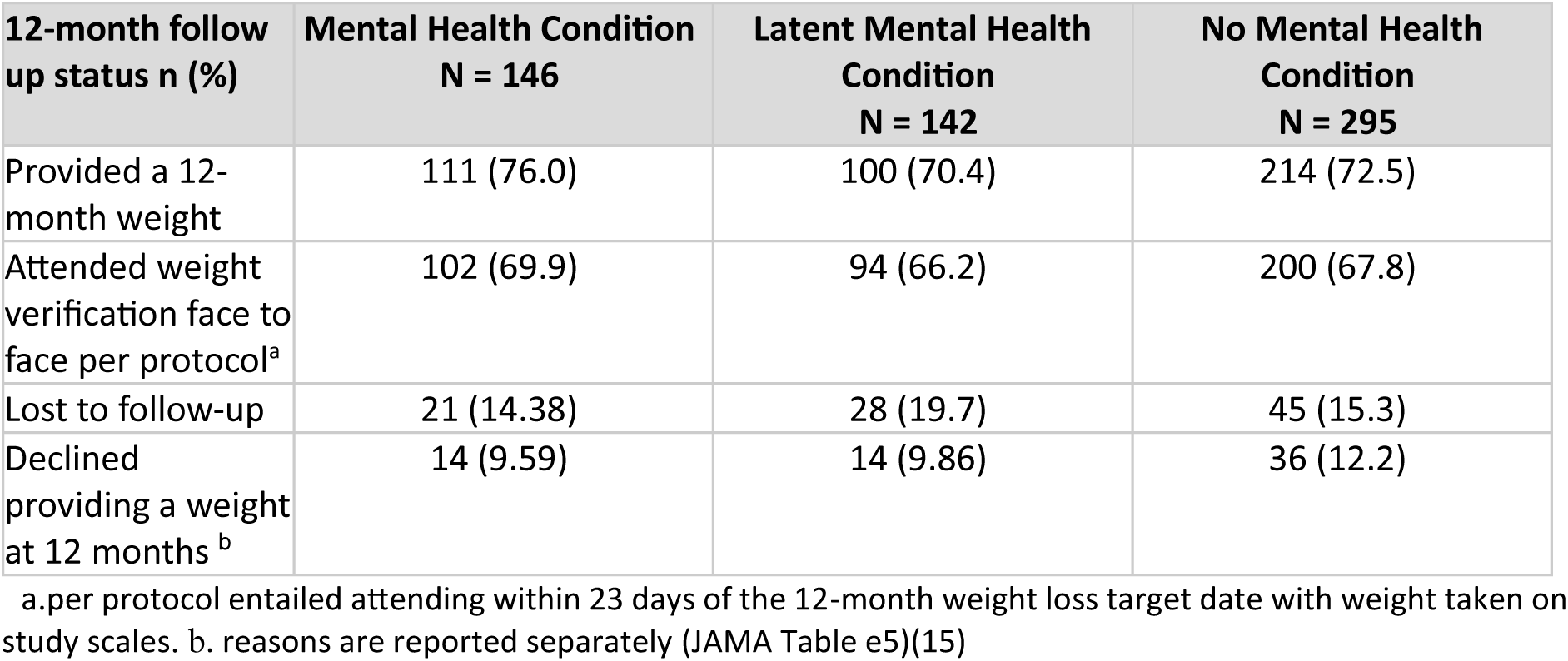
Retention of participants by baseline mental health category at 12-month weight assessment

| 12-month follow up status n (%) | Mental Health Condition<br>N = 146 | Latent Mental Health Condition<br>N = 142 | No Mental Health Condition<br>N = 295 |
| --- | --- | --- | --- |
| Provided a 12-month weight | 111 (76.0) | 100 (70.4) | 214 (72.5) |
| Attended weight verification face to face per protocol<sup>a</sup> | 102 (69.9) | 94 (66.2) | 200 (67.8) |
| Lost to follow-up | 21 (14.38) | 28 (19.7) | 45 (15.3) |
| Declined providing a weight at 12 months<sup>b</sup> | 14 (9.59) | 14 (9.86) | 36 (12.2) |

a. per protocol entailed attending within 23 days of the 12-month weight loss target date with weight taken on study scales. b. reasons are reported separately (JAMA Table e5)(15)

Although mental health categories were imbalanced, some differences were observed. Multiple long-term conditions were reported by 56.2% (n=82) men with a mental health condition 36% (n=107) with no mental health condition and 32.4 (n=46) with a latent mental health condition; 49.0% (n=71) with a mental health condition met the Office for National Statistics criteria for disability versus 20.5% (n=60) with no mental health condition and 24.1% (n=34) with a latent mental health condition; 12.8% (n=18) men with a mental health condition were not working due to disability compared to 2.1% (n=6) of those with no mental health condition and 3.5% (n=5) with a latent mental health condition; 22.4% men with a mental health condition reported being single versus 9.7% (n=28) with no mental health condition and 11.3% (n=16); with a latent mental health condition ;66.4% (n=97) with a mental health condition were living with a partner versus 80.3% (n=237) with no mental health conditions and 79.6% (n=113) with a latent mental health condition; 17.7% (n=25) with a mental health condition were finding it quite or very difficult financially versus 2.4% (n=7) with no mental health condition and 13.5% (n=19) with a latent mental health condition. Previous attempts at weight loss also differed: men with a latent mental health condition, a mental health condition and no mental health condition reported a median of 10 attempts, 7 attempts and 5 attempts respectively (Table 1; S1 Table).

There was overlap between mental health categories and living in disadvantaged areas in the trial population and in the qualitative sample (Figure 1, Table 1).

**Figure 1.**
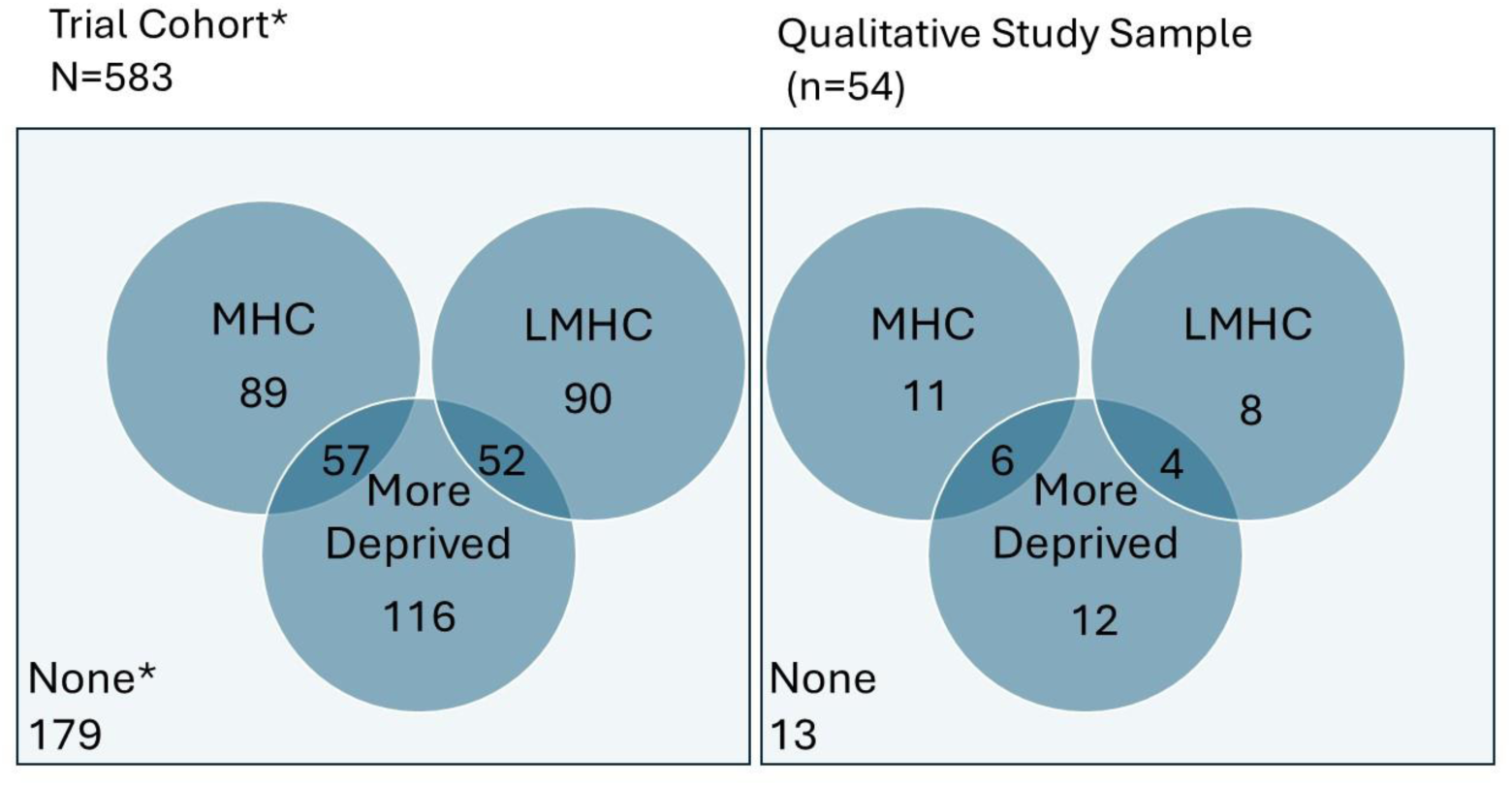
Participants in main trial and qualitative study by deprivation and mental health category. *Excludes 2 participants who could not be classified at baseline due to missing information. ‘More deprived’ refers to the two more deprived UK post-code quintiles for the Index of Multiple Deprivations.

### Retention

Of the total 585 trial participants, 426 (73.0%) provided a weight measurement at 12 months (Hoddinott, 2024). Of these, 76.0% (n=111) men with a mental health condition, 70.4% (n=100) men with a latent mental health condition and 72.5% (n=214) men with no mental health condition provided a weight at 12 months.

### Exploration of mental health and wellbeing outcome measures

The latent mental health condition category was identified by poorer mental health scores on either WSSQ, WEMWBS, PHQ-4 and/or EQ-5D-5L-AD. The contribution of each of these scores to identifying men in the latent mental health category is summarised in Table 3, with the number of outcome measures indicating poorer mental health reported in S1 Table.

**Table 3:** Number (%) of participants with mental health and wellbeing measure scores indicating a latent mental health condition by trial group.

| Outcome Measure | Texts + Incentive (N=50) | Texts Alone (N=48) | Control (N=44) |
| --- | --- | --- | --- |
| WSSQ | 36 (25.4) | 26 (18.3) | 25 (17.6) |
| WEMWBS | 27 (19.0) | 21 (14.8) | 24 (16.9) |
| PHQ-4 | 13 (9.2) | 18 (12.7) | 8 (5.6) |
| EQ-5D-5L-AD | 1 (0.7) | - | 1 (0.7) |

Weight stigma scores were high (42+) for 87 (61%) of the 142 latent mental health condition participants, with 72 (51%) reporting low wellbeing scores on WEMWBS (14–40). For men with a latent mental health condition, 90/142 (63.4%) participants were identified from scores on one outcome measure the majority of which were identified by weight stigma. 46 (32%) were identified from scores on two outcome measures and six (4.2%) from scores on three outcome measures (S1 Table).

For the 146 men with a mental health condition scores indicating poorer mental health are reported in S3 and S4 Tables. WSSQ and WEMWBS scores were the most common indicators of poorer mental health, with 62 men with no indicator, 40 had 1 indicator, 21 had 2 indicators, 15 had 3 indicators, 8 had 4 indicators.

The mental health and wellbeing outcomes for men with a mental health condition, a latent mental health condition and no mental health condition according to trial group are reported in Table 4.

**Table 4.** Mental health and wellbeing measures by mental health status and trial group.

| Variable –<br>mean<br>(SD); n | Baseline |  |  |  |  |  |  |  |  | 12 Months |  |  |  |  |  |  |  |  |
| --- | --- | --- | --- | --- | --- | --- | --- | --- | --- | --- | --- | --- | --- | --- | --- | --- | --- | --- |
|  | Texts + Incentives |  |  | Texts Alone |  |  | Control |  |  | Texts + Incentives |  |  | Texts Alone |  |  | Control |  |  |
|  | MHC<br>(n=51) | LMHC<br>(n=50) | No<br>MHC/L<br>MHC<br>(n=94) | MHC<br>(n=46) | LMHC<br>(n=48) | No<br>MHC/L<br>MHC<br>(n=99) | MHC<br>(n=49) | LMHC<br>(n=44) | No<br>MHC/<br>LMHC<br>(n=102) | MHC<br>(n=51) | LMHC<br>(n=50) | No<br>MHC/L<br>MHC<br>(n=94) | MHC<br>(n=46) | LMHC<br>(n=48) | No<br>MHC/<br>LMHC<br>(n=99) | MHC<br>(n=49) | LMHC<br>(n=44) | No<br>MHC/L<br>MHC<br>(n=102) |
| WEMWBS | 43.71<br>(9.79);<br>51 | 40.82<br>(7.81);<br>50 | 52.11<br>(6.67);<br>91 | 40.37<br>(9.49);<br>46 | 42.04<br>(6.64);<br>48 | 52.10<br>(6.41);<br>97 | 44.17<br>(7.10);<br>48 | 40.28<br>(7.80);<br>43 | 50.96<br>(5.98);<br>101 | 44.63<br>(10.22);<br>38 | 45.84<br>(8.87);<br>38 | 53.56<br>(8.20);<br>66 | 43.18<br>(9.25);<br>33 | 43.04<br>(6.51);<br>28 | 51.06<br>(8.78);<br>64 | 43.95<br>(9.00);<br>39 | 44.27<br>(9.09);<br>33 | 50.08<br>(9.66);<br>79 |
| PHQ4 | 4.03<br>(3.22);<br>30 | 3.22<br>(2.57);<br>41 | 1.01<br>(1.19);<br>69 | 4.46<br>(3.26);<br>37 | 3.86<br>(2.44);<br>35 | 0.90<br>(1.09);<br>68 | 4.00<br>(3.35);<br>32 | 2.78<br>(1.70);<br>32 | 1.05<br>(1.14);<br>78 | 3.39<br>(3.35);<br>36 | 2.63<br>(2.76);<br>38 | 1.32<br>(2.10);<br>65 | 4.03<br>(3.00);<br>32 | 3.04<br>(2.78);<br>27 | 1.55<br>(2.22);<br>62 | 3.26<br>(3.45);<br>39 | 2.88<br>(2.11);<br>32 | 1.36<br>(1.61);<br>78 |
| WSSQ | 36.89<br>(9.21);<br>50 | 43.32<br>(5.40);<br>50 | 31.02<br>(6.32);<br>93 | 38.50<br>(8.78);<br>46 | 40.95<br>(6.60);<br>48 | 30.19<br>(6.84);<br>98 | 37.54<br>(8.80);<br>48 | 41.53<br>(7.06);<br>44 | 31.81<br>(6.20);<br>99 | 34.84<br>(9.06);<br>37 | 39.07<br>(6.48);<br>38 | 30.60<br>(7.50);<br>66 | 36.70<br>(9.41);<br>33 | 37.90<br>(7.03);<br>28 | 29.27<br>(7.42);<br>66 | 39.32<br>(7.79);<br>40 | 38.61<br>(7.51);<br>33 | 30.94<br>(6.36);<br>79 |
| EQ-5D-5L | 0.64<br>(0.26);<br>51 | 0.70<br>(0.22);<br>50 | 0.79<br>(0.20);<br>93 | 0.62<br>(0.24);<br>45 | 0.70<br>(0.22);<br>48 | 0.82<br>(0.20);<br>99 | 0.64<br>(0.24);<br>49 | 0.67<br>(0.21);<br>43 | 0.78<br>(0.16);<br>99 | 0.66<br>(0.29);<br>38 | 0.72<br>(0.23);<br>37 | 0.84<br>(0.19);<br>67 | 0.60<br>(0.28);<br>33 | 0.71<br>(0.24);<br>28 | 0.78<br>(0.24);<br>66 | 0.71<br>(0.23);<br>40 | 0.73<br>(0.19);<br>33 | 0.82<br>(0.16);<br>76 |
| EQ-5D-VAS | 57.20<br>(18.34);<br>50 | 56.28<br>(17.47);<br>50 | 66.67<br>(18.30);<br>94 | 58.05<br>(22.07);<br>43 | 57.68<br>(17.69);<br>47 | 67.59<br>(16.93);<br>96 | 59.77<br>(19.40);<br>48 | 55.44<br>(17.30);<br>43 | 66.33<br>(18.64);<br>102 | 67.67<br>(17.67);<br>36 | 69.00<br>(17.29);<br>38 | 71.40<br>(17.80);<br>67 | 62.29<br>(20.23);<br>31 | 65.63<br>(19.11);<br>27 | 73.35<br>(18.75);<br>63 | 62.10<br>(17.68);<br>40 | 62.94<br>(14.85);<br>33 | 68.87<br>(16.35);<br>75 |
| EQ-5D-5L-AD | 2.55<br>(1.05);<br>51 | 2.02<br>(0.87);<br>50 | 1.37<br>(0.62);<br>93 | 2.57<br>(1.03);<br>46 | 2.19<br>(0.70);<br>48 | 1.27<br>(0.53);<br>99 | 2.69<br>(0.98);<br>49 | 1.91<br>(0.94);<br>44 | 1.50<br>(0.66);<br>101 | 2.53<br>(1.03);<br>38 | 2.03<br>(1.20);<br>38 | 1.36<br>(0.64);<br>67 | 2.64<br>(1.06);<br>33 | 2.07<br>(0.86);<br>28 | 1.34<br>(0.66);<br>67 | 2.33<br>(1.10);<br>40 | 1.88<br>(0.74);<br>33 | 1.57<br>(0.79);<br>77 |

Abbreviations: MHC-Mental Health Condition, LMHC-Latent Mental Health condition.

Baseline scores for all six mental health, wellbeing and quality of life measures were worse for men with a mental health condition and men with a latent mental health condition compared to men who had no mental health or latent mental health condition.

Quality of life (EQ-5D VAS) improved between baseline and 12 months regardless of mental health category across all trial groups. However, changes in quality of life (EQ-5D-5L) were inconsistent, as for the trial population overall(15). Mental wellbeing (WEMWBS) improved at 12 months for men with a mental health condition in both intervention groups and for men with a latent mental health condition across all trial groups. For men with no mental health condition, mental wellbeing improved at 12 months for men in the texts and financial incentives group. Weight stigma (WSSQ) improved for both intervention groups, regardless of mental health category. Anxiety and Depression (PHQ-4) improved at 12 months for men with a mental health condition across all trial groups and for men with a latent mental health condition in both intervention groups. Anxiety and Depression (PHQ-4) was worse at 12 months for men with no mental health condition across all trial groups. A summary of all differences observed between baseline and 12 months are provided in S5 Table. These require cautious interpretation due to the imbalance for the three mental health categories.

### Qualitative findings

Interviews were held with 54 men across the three trial centres between September 2022 and May 2023 (Table 5). They lasted on average 40 minutes (range: 13-81 minutes). Participants were aged between 24 and 81 years old (Mean: 55.6 years) and were mainly of white ethnic background. Of those interviewed, 17 self-reported ever having a mental health condition, 12 were identified as having a potential latent mental health condition, and 25 men had no mental health condition at baseline. 28 participants had met one or more of their weight loss targets over the course of the 12-month programme (Table 5).

**Table 5.** Characteristics of men participating in interviews

| Characteristic | Texts Alone | Texts + Incentive | Total |
| --- | --- | --- | --- |
| **Mental Health Status at baseline** |  |  |  |
| Mental Health Condition | 3 | 14 | 17 |
| Latent Mental Health Condition | 4 | 8 | 12 |
| No Mental Health Condition | 17 | 8 | 25 |
| **Postcode-Based Deprivation Status at baseline** |  |  |  |
| Less Deprived (IMD 3,4,5) | 13 | 18 | 31 |
| More Deprived (IMD 1,2) | 10 | 12 | 22 |
| **Weight Loss Outcome** |  |  |  |
| Met one or more targets | 7 | 21 | 28 |
| Met no targets | 17 | 9 | 26 |

Abbreviations: IMD-Index Multiple Deprivation

To explore the impact of mental health on men’s experiences of the trial, the findings below present the accounts of men ever living with a self-reported mental health condition or a latent mental health condition. We start by detailing how men in these groups talked about the impact of mental health and wellbeing on their ability to make changes and lose weight. We then present the patterns observed relating to their experiences of the core intervention components, namely the financial incentives, text messages, and weight loss targets. Where appropriate, fieldworker notes detailing summaries of participants’ responses to the harms and benefit question have been integrated with the interview data. Responses taken from harms and benefits data are highlighted throughout the text in italics and labelled PR.

### Mental health in the context of weight loss

Participants in each of the mental health categories spoke of their experience of mental health while taking part in the trial. Men with a mental health condition described common mental health conditions, including anxiety and depression, stress, and more severe mental health conditions such as binge eating disorder and bipolar disorder. Men often spoke of their mental health and wellbeing co-existing alongside long-term physical health problems, such as arthritis, chronic obstructive pulmonary disease, hypertension and sleep apnoea; others described more acute problems such as undergoing surgery or sustaining injuries during their time in the trial.

Men living with a mental health condition occasionally talked about the negative impact of poorer mental health on their ability to lose weight. Within the harms and benefits data some men attributed weight gain to taking antidepressants, as one participant noted ‘*mental health remains poor and has meant that weight loss just isn’t a priority*’ (PR, Texts with Incentives, Less Deprived, MHC). It was also apparent that some men faced multiple stressors and that the context in which they were experiencing them, exacerbated them further: ‘*Still trying to move forward from [parent’s] death last year, very difficult time. Lost his job just before Christmas which has been financially very hard with the cost-of-living crisis compounding things’* (PR, Texts Alone, More Deprived, MHC).

Overall, however, there was limited evidence of poorer mental health hindering weight loss and in some cases, those who had ongoing difficulties with their mental health were able to meet at least some of their weight loss targets, even exceeding them. Where mental health was described during the interviews as a barrier to behaviour change, this was often related to a specific period of poorer mental health during the trial.

> *Well personally I suffer quite bad depression. So when I do get depressed I don’t want to leave the room…Eating, even socialising, anxiousness a lot…Cos and I also eat too much…I*
>
> *feel as if I’m entitled to overdose [on food] and really…when it does hit, it can hit quite hard*.
>
> (Interview, Texts with Incentives, More Deprived, Met no Targets, MHC)

There was a tendency among men with a mental health condition to talk about the social barriers they faced to losing weight, such as having limited support, family and work circumstances, with the Christmas period being particularly difficult to manage, in addition to physical illness and injury.

> *I don’t have any kind of support group round about me. I don’t have family or friends. There’s me and four walls. Whilst there are benefits to that, I don’t have anybody clipping me around the ears when I back slide.* (Interview, Texts with Incentives, More Deprived, Met one or more Targets, MHC)

For men identified as having a latent mental health condition there were some specific discussions during interviews about mental health. There were descriptions of experiencing periods of depression during the trial. However, these men more commonly spoke about the life stressors they had experienced, such as work stress, caring for others, the cost-of-living crisis and societal changes (e.g. lockdowns) due to the COVID-19 pandemic as well as on health.

For some men with a latent mental health condition, periods of time where life was stressful sometimes impacted their ability to engage with behaviour change and therefore meet their weight loss targets. For example, during the interviews ‘emotional eating’ was commonly discussed and in the harms data it was noted alcohol was being used “*as a crutch*” to deal with a bereavement (PR, Texts with Incentives, LMHC). However, experiencing stress did not always prevent behaviour change and weight loss, with some still able to make progress.

Men with a latent mental health condition often spoke about injuries, illnesses, as well as longer-term physical health harms they experienced over the course of the trial rather than experiencing specific mental health problems. The inter-related nature of physical health and mental health was often described and is reported elsewhere(29).

> *That [knee injury] was definitely a huge negative which in turn had a psychological impact as well because I felt frustrated, disappointed, disheartened at times. It then meant that other areas that could have contributed such as healthier eating…Just lost more motivation to try that type of food because of the limitations on my mobility* (Interview, Texts Alone, More Deprived, Met no Targets, LMHC)

This was also evident in the harms data as men with a mental health condition or a latent mental health condition, who had lost some weight, reported improved physical health (e.g. more energy and mobility, less hip and knee pain, improved breathing, reverting from type 2 diabetic to pre-diabetic blood levels), being more active, returning to previous hobbies (walking, playing golf and football) and being more sociable. Whilst men did not always directly associate these changes to improved mental health, there was some suggestion that they were experiencing less pain and a better quality of life. Some men were noted as enjoying the changes they had made and feeling better mentally.

> *He said that he hadn’t been expecting the boost in mood that he’s experienced since taking part in the trial. There have been other things that have contributed but Game of Stones has been a factor. Losing weight and having goals has been very helpful to him* (PR, Texts with Incentives, MHC)

### The value of financial incentives

Men regardless of mental health category had mixed views regarding the acceptability of financial incentives with men voicing both positive and negative perspectives. Some men saw the incentive as motivational, supporting their own weight loss, or could see how it would be motivating for other people. Although incentives were not explicitly advertised men talked about how the potential to receive money was sometimes the reason they had signed up for the study. In the harms data it had been noted that the ‘*incentive had been a catalyst for change. A way to make money and feel healthier’* (PR, Texts with incentives, More Deprived, LMHC). For some, the money was a necessity providing a financial boost, particularly in the context of limited income or wealth.

> *I was fortunate to get the one with the potential for £400 at the end of the year. So that was a huge incentive because I’m part…I’m retired, but I only get a part pension, so any extra funds was gratefully received* (Interview, Texts with Incentives, Less Deprived, Met All Targets, MHC)

Men in the texts and incentives group who were in favour of financial incentives, perceiving them as a good or sensible idea, generally had met all or most of their targets. There was sometimes a reluctance to or discomfort in acknowledging the role incentives played despite indicating their value and benefit, this appeared related to the negative perceptions around incentives. Some suggested they would have tried to lose weight without the incentive, that health was the real focus or that the incentive was an added bonus rather than the primary motivating factor.

> *I was always focused solely on losing weight and if I hit the targets and got the full amount then that would have been fantastic but that was never the kind of most important factor to me. The losing of the weight was more reward than the money could have been* (Interview, Texts with Incentives, Less Deprived, Met Some Targets, MHC).

Some men while initially having a positive view of the financial incentives found they were not motivating enough in their circumstances.

> *I thought it was a great idea, I just thought it was money in the bank, you know, thinking I thought if I really focus here, I could lose 10 pounds not a problem – but it was a problem (laughing). It’s not easy, the offer’s’ great, it just seems great on paper but not into practice, it’s just not a simple process but I can see it might work for some, and I thought it would work for me, but it didn’t* (Interview, Texts with Incentives, Less Deprived, Met no targets, LMHC).

Whereas others said that incentives did not motivate them at all and/or were not required for motivation. A few men described becoming demotivated by the incentive due to their limited success with weight loss. Some found their inability to attain the incentive became a source of embarrassment and on occasion a barrier to behaviour change. There was a belief that people should be internally motivated, for example by their own self-worth, and that personal weight loss or health gains should be enough, or that external factors such as family should be a principal motivator and not incentives. This appeared related to an understanding that weight is attributable to personal responsibility. Therefore, an intervention which offered money to lose weight was perceived to have a moral implication. Some thought that public acceptability of incentives would be low and that the money could be better spent elsewhere. Discomfort with the financial incentives was sometimes related to men’s sense that they were living more comfortably and did not require the money. Some of these men were from less deprived areas.

> *…I consider myself comfortable. I have a comfortable lifestyle. There’s probably people out there who are a lot worse off than me. Being given the money, being offered the money, yeah it felt a little bit guilty, a little bit ‘oh what will I do with this’ just uncomfortable with it. That was at the start of it. I don’t believe I would have performed any differently if I was not offered money* (Interview, Texts with Incentives, Less Deprived, Met All Targets, LMHC)

However, concerns were also raised by those with more severe mental health problems. One participant described how the possibility of obtaining any incentive would cause harm to his mental health and wellbeing. The stress he experienced of that potential outcome led to disengagement.

> *Because of my illness, I don’t want money around me, I **can’t** have money around me…part of the reason why I didn’t use it [the intervention] well, or I didn’t use it to its full potential you know, when I first went and I lost it down, the lady said you’ve now earned such as such money you know and I’m going - shit, this for my mental health is not a good thing. So that’s why I would say more than likely I stepped away from it.* (Interview, Texts with Incentives, More Deprived, Met some targets, MHC)

### The value of text messages

Men across all mental health categories held mixed views on the content and motivational nature of the text messages. On the one hand men described finding the text content valuable, in some cases supporting their behaviour change efforts, weight loss and missed the texts when they ended. On the other hand, some men disengaged and disliked the text messages. During interviews some men described how the frequency or timing of delivery and/or content led to behaviour change directly, such as providing them with information which helped them to adopt practical strategies such as standing rather than sitting, batch cooking or substituting one food for another, or led them toward resources and support that could assist them with changes.

***…****what I had been doing was binge eating at night and making plans to binge eat outside of the house and hiding food and this kind of stuff. So I’ve been able to identify those kind of negative behaviours and put in place processes to try and cut them out, so that has helped. And that came from the texts … So I think that’s one positive of it and that is probably something that will be key going forward for any kind of sustainable weight loss.* (Interview, Texts with Incentives, More Deprived, Met Some Targets, MHC)

The harms and benefits data also indicated that the texts had highlighted to men what they could change and how they could do this: *Texts are really helping to pinpoint areas that I could work on to improve my weight loss* (PR, Texts Alone, Less Deprived, LMHC). *Found text messages have increased awareness around food, including portion size* (PR, Texts Alone, Less Deprived, LMHC). There was a strong indication that men had found the texts served as a frequent prompt and functioned as a reminder to reinforce their desire to lose weight.

> *Some days, it depended what was going on, some days it was lovely, it was kind of a continual reminder you know, Game of Stones…Other days you could see me looking (makes noise) but you know they were either a pick-me-up, a pat on the back, or an itch that you just couldn’t reach, so no matter what mood I was in it was there, you know, so it was either very much welcome, or a gentle reminder, or it really annoyed, you know…* (Interview, Texts with Incentives, More Deprived, Met some Targets, MHC)

This was also evident in the harms and benefits data, where a fieldworker captured a participant describing how the texts had reminded him that he was trying to lose weight: *how the text messages have made him more focused and conscious of food choices and overall health behaviours. He said that the texts don’t guilt trip you into anything but are a good reminder of what you should be doing* (PR, Texts with incentives, Less Deprived, LMHC)

There was also a perception across interviews and in the harms and benefits data that the texts provided a supportive network and reminded men that they were not alone in struggling to lose weight. For some men, this idea was strengthened by their belief that the texts, and therefore ideas and suggestions about weight loss, were coming directly from other men in the trial. *So it was it was giving me that feeling that I’m still within the group… the problems I’m having with the weight maintenance, other people are having the same thing, the same problems, the same doubts, the same failures… It was always kind of like looking for brothers in misery (*Interview, Texts Alone, Less Deprived, Met No Targets, LMHC)

However, other men described at least some of the texts as monotonous, ‘nothing new’, irrelevant or ‘not for them’. This was also evident in the harms and benefits data where it was noted that the texts did not resonate with some men. Those holding such views questioned whether the texts offered any valuable insight to motivate or initiate change. For these men, even engagement with the texts, such as reading them regularly, failed to support behaviour change. Those who shared this viewpoint were less likely to have met their weight loss targets than those who spoke about the motivating nature of the texts.

> *And it didn’t really [help], I mean, it was all down to me. You know there was alright the daily message that came through, but as I say, it wasn’t something that made me stand up and say, ‘yeah, interesting. I’ll do this. I’ll try that and…’ Whether it’s just me I don’t know.* (Interview, Texts with Incentives, Less Deprived, Met no Targets, MHC)

Some men expected the texts to provide motivation, and these expectations were not met. For them, the frequent contact through messages did not elicit the sense of support they required and sometimes represented more of a burden or stressor, leading to disengagement.

> *And what the study didn’t do, obviously it wasn’t designed to which was absolutely fine, was to provide that support, that thicker more tangible, more in-depth support I needed at that point in time. What that meant was, the group I was in received text messages on a daily basis, but actually that was becoming more of a burden and more of a negative impact*.
>
> *These messages were coming through but actually I didn’t know what direction to take with the weight loss I was wanting to do* (Interview, Texts with Incentives, More Deprived, Met Some Targets, MHC)

### Weight loss targets - a realistic or unrealistic goal

Weight loss goals were generally regarded as motivational and helpful for the purpose of maintaining focus, monitoring progress and for accountability.

> *…it [weight loss targets] was something to aim for and I don’t really weigh myself, but I was weighing myself every 10 days or so, so that helped me focus, maybe gave me a bit of push to check my weight whereas beforehand I wouldn’t have bothered my backside*. (Interview, Texts with Incentives, Less Deprived, Met No Targets, LMHC)

However, there were suggestions that having targets or not meeting targets could be demotivating and problematic. In some cases, having weight loss goals sometimes became a barrier to behaviour change, shifting the perspective on what was achievable. These men reported that achieving the weight loss targets was more challenging than they had initially envisaged. Personal circumstances or having health problems were often cited reasons for not meeting the programme targets or their own personal targets. Some men also suggested their inability to meet the weight loss targets was related to the self-care ethos of Game of Stones. Their inaction was for example attributed to not having group-based support or having targets which were too long-term. Although, men with a mental health condition often described the targets as reasonable, some also talked about them as not challenging enough, too easy or set over too long a period. The expected weight loss during the trial was high (S1 Table) and for some men their expected weight loss could be considered unrealistic. Improving health was important to men. However, some did not appear to make the connection between meeting weight loss goals and improving health. The weight loss goals participants were expectant of therefore seemed somewhat arbitrary and detached from the programme.

> *It’s sort of in my mind I was minimising the effectiveness of the study for me. You know what I mean? ‘No, this [weight loss target] not what you really want because you want 10 stone off’ and I went back in and I was heavier than when I started* (Interview, Texts Alone, Less Deprived, Met no targets, MHC)

Some men expressed a negative perception of their weight loss, indicating that they should have done better, even when they had met some or all of the programme targets. These men talked about what they should have achieved or wanted to achieve rather than the actual outcomes. This appeared related to personal responsibility as these men attributed their inability to meet the targets, they or the programme had set, to their own personal failings rather than to other factors.

> *5% seems like nothing, and I guess it’s a very small number. It doesn’t sound much, and I never really thought too much of actually how much weight that had to be. It was whenever I started trying to lose the weight and I got there but I only just kind of got there. I could have done better, but I didn’t do better and that all was down to me and I began to realise that…* (Interview, Texts with Incentives, More Deprived, Met all Targets, MHC)

## Discussion

This mixed methods study explored and triangulated trial data with men’s experiences of mental health conditions and obesity during a randomised controlled trial of text messages with or without financial incentives. Health inequalities were evident for men reporting ever having a mental health condition; more were single, had a disability, were living with multiple long-term conditions and under financial strain compared to men with no mental health conditions. Yet trial retention for the primary outcome of percentage weight change at 12 months was higher than trial average for men with a mental health condition and lower for men with a latent mental health condition (baseline scores indicating poor mental health but did not self-report a mental health condition).

The three mental health categories did not moderate the effectiveness for percentage weight lost at 12 months for either texts with incentives or texts alone(27). The qualitative findings mirrored these results, with both interventions appealing to and helping some men but not others. The narratives of men regardless of mental health category provided evidence of a high degree of commonality in terms of experiencing life stressors, physical health problems, and periods of mental ill health which may or may not have previously been diagnosed. The programme was also delivered during a period of unprecedented challenges with the COVID pandemic and cost-of-living crisis. These inter-related difficulties meant that men’s lives and experiences were complex. Common mental health problems were discussed, such as stress, anxiety and depression. These were experienced in different ways, often unique to the individual, and could be episodic, recurrent, cyclical or ongoing. Men who had lived experience did not always talk of their mental health as a barrier to behaviour change. In some cases, these men were highly engaged and successfully met weight loss targets. Whereas those living through mental health struggles during the trial described how this challenged behaviour change and weight loss.

The enrolment and retention of a high proportion of trial participants who reported ever been told they had a mental health condition or a latent health condition demonstrates the broad appeal of Game of Stones. It is also unusual for weight management trials to recruit men from lower-socio-economic groups(12, 33, 34). This level of continuing engagement is contrary to a previous secondary analyses on mental health from the WRAP trial(33) where participants who reported poorer baseline mental health were less likely to attend programme sessions, engage with resources, and attend study assessments. However, a previous cluster trial of a 12-month diabetes prevention programme with financial incentives which enrolled predominantly low-income women and 40% reported a previous mental health condition, demonstrated that mental health did not affect retention(35). Acceptability of financial incentives for behaviour change continues to be problematic for some but not others, as found in our feasibility study and was not determined by mental health category(20). Financial incentives made a difference, particularly to some men on low incomes, and the acceptability is likely to grow as the evidence of effectiveness increases(36).

Identifying men at baseline who may have an undiagnosed mental health condition or are potentially at risk of developing one (latent mental health) is, as far as we are aware, a unique approach within behaviour change trials. It was undertaken to understand why fewer men report mental health problems compared to women in all age categories(13), and why men with mental health problems have lower engagement with lifestyle interventions than men without(12). Men with latent mental health conditions were usually identified by high scores for weight stigma or low wellbeing scores, with few identified by brief anxiety and depression measures at baseline. Baseline scores for all six mental health, wellbeing and quality of life measures were worse for men in the mental health and latent mental health categories compared to men with no mental health condition. Small improvements in scores were observed in wellbeing, weight stigma and PHQ-4 anxiety and depression scores for men in the mental health and latent mental health categories, for both texts with incentives and texts alone, but EQ-5D-5L-AD and EQ-5D-5L scores were inconsistent. Our findings illustrate that it remains unclear which is the most appropriate and valid mental health and wellbeing measure to detect changes in responses for men with obesity recruited from community or primary care settings.

### Strengths and Limitations

A strength of this study is the unusual and underserved trial population for investigating mental health in a behavioural weight management trial for men, which included lower socio-economic groups, men with disability and multiple long-term conditions. Importantly the findings begin to fill a recognised gap in the evidence for gender-tailored interventions for men with obesity and mental health conditions(12). The identification of a latent mental health condition category is unique and provides valuable new insights. The mixed methods approach with triangulation of quantitative and qualitative data illustrates the potential for exploring underserved sub-groups in a trial.

There are several limitations. Generalisability to men from diverse cultures, with low literacy or without regular access to a mobile phone is unknown. This study is exploratory and hypothesis generating only, indicating where further research might be of value. The PHQ-4 data collection commenced late, and therefore has more missing data at baseline compared to the other secondary outcomes. There was some missing data for several baseline characteristics which were evenly distributed across trial groups. The imbalance of mental health category size is a further limitation.

Therefore, a very cautious interpretation is required when comparing outcomes by mental health category. The interventions were not designed to improve mental health outcomes, unlike the Australian online men-only SHED-IT trial(37). Using several measures and completing the trial during the Covid-19 pandemic lockdowns, which were a stressful time, may have impacted scores at baseline and 12 months which provide a single snapshot of mental health, wellbeing and quality of life.

## Conclusions

Trial retention for men ever diagnosed with a mental health condition was above average for the trial, whereas those in the latent mental health category was lower. The measures for mental health and wellbeing showed different patterns of results according to mental health category, illustrating a continued uncertainty about the most suitable outcomes for men participating in this behavioural weight management trial. Regardless of mental health category, the Game of Stones text message with or without financial incentives interventions helped some men to lose weight and not others. Findings suggest that some men were able to overcome difficulties related to stress and mental health problems and still attain their weight loss goals, while others struggled.

## Data Availability

The data collected for the study, including individual patient data and a data dictionary defining each field in the data set will be made available to others. The participant data will be de-identified and will comply with the ethical and regulatory approvals for the study. Requests for access to data can be sent by email to and will be considered by the study team.

## Acknowledgements

The authors would like to thank Kathryn Machray^1^ who undertook some interviews, Abraham Getaneh^2^, Dwayne Bower^2^ and Marjon van der Pol^2^ of the Project Management Team, and all the trial participants who made this research possible. We thank the general practices, clinical research networks, community workers and local stakeholders who advertised the trial; the people who have contributed public and patient involvement to improve the research design, materials and conduct of this study, particularly during the Covid-19 pandemic. The trial fieldworkers were outstanding in their work on recruitment and data collection: Kathryn Machray^1^, Norelle Calder-McPhee^1^, Clare Jess^3^, Christina O’Neill^3^, Angela Mullan^3^, Hilary Taylor^4^, Jack Brazier^4^, and all the students who assisted. We thank Matthew McDonald^5^ and other team members from the feasibility trial who shared their experiences of recruiting and collecting data. The contributions of the Men’s Health Forum (GB and Ireland) since 2010 have been invaluable for the design and conduct of this study; our thanks to Martin Tod^6^, Jim Pollard^6^, Colin Fowler^7^, Paula Caroll^8^ and Michael McKeon^9^.

We would also like to thank the independent members of the Trial Steering Committee: Prof. Edmund Juszczak (Chair)^10^, Prof. Emma Frew^11^, Mr David Gardner (lay member and Chairman of Scottish Men’s Sheds)^12^, Mr Graham Jameson (lay member and participant in the Football Fans In Training trial), and Prof. Kate Jolly^11^ for their oversight and guidance. Mr Gardner and Mr Graham received compensation for their contribution. We acknowledge the contributions to the trial protocol of people who are no longer involved with the study: Andrew Elders^13^ and Beatriz Goulao^14^ for statistics contributions, Fiona M Harris^15^ for process evaluation contributions.

We are grateful for the technical administrative support and database/website development of Mark Forrest^14^, Connor Keegan^14^ and the team at the Centre for Healthcare Randomised Trials^14^. We extend thanks to Claire Jones^16^, Jack Gilmore^16^, Ross Teviotdale^16^, and Keith Milburn^16^ who developed the participant tracker software and delivered the text intervention. We acknowledge the earlier work on text interventions of Professor IK Crombie^17^.

1. University of Stirling, UK.
2. Health Economics Research Unit, University of Aberdeen, UK.
3. Queens University Belfast, UK.
4. University of Bristol, UK.
5. Curtin University, Australia.
6. Men’s Health Forum, London, UK.
7. Men’s Health Forum in Ireland, Dublin, Ireland.
8. South East Technological University, Waterford, Ireland.
9. Dublin City University, Ireland.
10. University of Nottingham, UK.
11. University of Birmingham, UK.
12. Scottish Men’s Sheds Association, Banchory, UK.
13. Glasgow Caledonian University, UK.
14. Centre for Healthcare Randomised Trials, University of Aberdeen, UK.
15. University of the West of Scotland, UK.
16. Health Informatics Centre, University of Dundee, UK.
17. University of Dundee, UK.

## Author Contributions

**Concept and design**: Hoddinott, O’Dolan, Macaulay, Dombrowski, Swingler, Avenell, Gray, Hunt, Kee, McKinley, Torrens, Turner, MacLennan. **Acquisition, analysis, or interpretation of data**: Hoddinott, O’Dolan, Macaulay, Dombrowski, Swingler, Cotton, Avenell, Hunt, Kee, MacLean, McKinley, Torrens, Turner, MacLennan. **Drafting of the manuscript**: Torrens, Turner, Macaulay, O’Dolan, Hoddinott, Swingler, MacLennan. **Critical review of the manuscript for important intellectual content**: All authors. **Statistical analysis:** Swingler, MacLennan. **Obtained funding**: Hoddinott, Dombrowski, Avenell, Gray, Hunt, Kee, McKinley, Turner, MacLennan. **Administrative, technical, or material support**: Hoddinott, O’Dolan, Macaulay, Cotton. Supervision: Hoddinott, McKinley, Turner, MacLennan.

## Conflict of Interest Disclosures

Dr Hoddinott reported receiving grants from National Institute for Health Research (NIHR), and the Chief Scientist Office (CSO), Scotland, during the conduct of the study and serving as chair or member of Independent Trial Steering Committees unrelated to weight management trials; being a member of the NIHR School for Primary Care Research Funding panel. Dr Dombrowski reported receiving grants from the NIHR during the conduct of the study. Mr Swingler reported receiving grants from NIHR during the conduct of the study. Dr Cotton reported receiving grants from NIHR HTA (grant funding to institution) during the conduct of the study. Dr Avenell reported receiving grants from National Institute for Health and Care Research funding project in submission during the conduct of the study. Dr Hunt reported receiving grants from NIHR, CSO, the Australian Heart Foundation, and the Department of Health, Australia, during the conduct of the study; and serving as chair of the Health Improvement, Protection and Services Committee of the Chief Scientist Office. Dr MacLean reported receiving grants from the NIHR during the conduct of the study. Dr McKinley reported receiving grants from NIHR during the conduct of the study. Ms Torrens reported receiving grants from NIHR during the conduct of the study. Dr Turner reported receiving grants from NIHR during the conduct of the study and having had served as a member of the NIHR Health Technology Assessment (HTA) commissioning board, December 2017 to September 2020. Mr MacLennan reported receiving grants from the NIHR during the conduct of the study. No other disclosures were reported.

## Supplementary Files

S1 Table All baseline characteristics according to mental health status

S2 Table. Number (%) of mental health and wellbeing outcomes identifying a latent mental health condition in participants.

S3 Table. Number of mental health and wellbeing baseline outcomes indicative of a latent mental health condition in participants (N, %) with a mental health condition by trial group.

S4 Table. Number (%) of participants with a mental health condition (n=146) with mental health and wellbeing outcome measure scores indicative of a latent mental health condition by trial group.

S5. Topic Guide

## Notes

### Competing Interest Statement

The authors have declared no competing interest.

### Clinical Trial

ISRCTN91974895

### Funding Statement

This trial was funded by the National Institute for Health and Care Research (NIHR), UK (Ref: NIHR 129703) using UK aid from the UK Government to support global health research. The research team were invited to apply for additional funding from NIHR in 2021 to investigate UK policy priority areas: mental health conditions and multiple long-term conditions within the Game of Stones trial. The views expressed in this publication are those of the authors and not necessarily those of the NIHR or the UK government. This project was supported by NHS Bristol, North Somerset and South Gloucestershire Integrated Care Board; NHS Greater Glasgow and Clyde; NRS Primary Care Network and HSC R&D Division of the Public Health Agency [HSC R&D Award Reference PHR Project: NIHR129703].

