## Supplementary Tables S1-S5 for "Mental health and weight loss in men: an exploratory mixed methods study of the Game of Stones trial"

### Supplementary Files

**S1 Table All baseline characteristics according to mental health status.**

| | No Mental Health Condition n=295 | Mental Health Condition n=146 | Latent Mental Health Condition n=142 |
|---|---|---|---|
| **Age* - mean (SD); n** | 53.4 (13.4); 295 | 47.5 (13.0); 145 | 48.4 (12.2); 142 |
| ≥18-<25 | 3 (1.0) | - | 3 (2.1) |
| ≥25-<45 | 76 (25.8) | 67 (46.2) | 48 (33.8) |
| ≥45-<65 | 152 (51.5) | 65 (44.8) | 76 (53.5) |
| ≥65-<75 | 48 (16.3) | 10 (6.9) | 15 (10.6) |
| ≥75 | 16 (5.4) | 3 (2.1) | - |
| **Deprivation Category - n (%)** | **n=293** | **n=144** | **n=142** |
| Most deprived | 67 (22.9) | 37 (25.7) | 29 (20.4) |
| More deprived | 49 (16.7) | 20 (13.9) | 23 (16.2) |
| Deprived | 38 (13.0) | 26 (18.1) | 23 (16.2) |
| Less deprived | 55 (18.8) | 26 (18.1) | 29 (20.4) |
| Least deprived | 84 (28.7) | 35 (24.3) | 38 (26.8) |
| **Ethnic Group* - n (%)** | **n=284** | **n=139** | **n=141** |
| White | 264 (93.0) | 133 (95.7) | 128 (90.8) |
| Mixed/ multiple ethnic groups | 4 (1.4) | - | 2 (1.4) |
| Asian/ Asian British | 8 (2.8) | 1 (0.7) | 2 (1.4) |
| Black/ African/ Caribbean/ Black British | 7 (2.5) | - | 2 (1.4) |
| Other | 1 (0.4) | 3 (2.2) | 4 (2.8) |
| Prefer not to say | - | 2 (1.4) | 3 (2.1) |
| **Relationship status* - n (%)** | **n=290** | **n=143** | **n=141** |
| Single (never married; never in a civil partnership) | 28 (9.7) | 32 (22.4) | 16 (11.3) |
| Co-habiting | 43 (14.8) | 29 (20.3) | 24 (17.0) |
| Married / civil partnership | 198 (68.3) | 69 (48.3) | 88 (62.4) |
| Separated | 6 (2.1) | 6 (4.2) | 2 (1.4) |
| Divorced | 7 (2.4) | 5 (3.5) | 7 (5.0) |
| Widowed | 4 (1.4) | 1 (0.7) | 1 (0.7) |
| Prefer not to say | 4 (1.4) | 1 (0.7) | 3 (2.1) |
| **Comorbidities*^ - n (%)** | **n=295** | **n=146** | **n=142** |
| No Comorbidities* | 111 (37.6) | - | 56 (39.4) |
| One Comorbidity* | 77 (26.1) | 64 (43.8) | 40 (28.2) |
| Multiple Comorbidities* | 107 (36.3) | 82 (56.2) | 46 (32.4) |
| Multiple comorbidities including self-reported diabetes | 47 (15.9) | 23 (15.8) | 20 (14.1) |
| High Blood Pressure | 135 (45.8) | 61 (41.8) | 66 (46.5) |
| Arthritis | 75 (25.4) | 35 (24.0) | 32 (22.5) |
| Diabetes | 59 (20.0) | 23 (15.8) | 22 (15.5) |
| Heart condition such as angina or atrial fibrillation | 51 (17.3) | 16 (11.0) | 24 (16.9) |
| Stroke (including TIA) | 10 (3.4) | 5 (3.4) | 5 (3.5) |
| Cancer | 13 (4.4) | - | 6 (4.2) |
| ONS-defined Disability* | **n=292** | **n=145** | **n=141** |

| | No Mental Health Condition n=295 | Mental Health Condition n=146 | Latent Mental Health Condition n=142 |
|---|---|---|---|
| Yes | 60 (20.5) | 71 (49.0) | 34 (24.1) |
| **Perceived wealth* - mean (SD); n** | **n=278** | **n=140** | **n=132** |
| Perceives to live in relatively wealthy neighbourhood (0-100, Strongly disagree) | 56.4 (28.6); 278 | 54.1 (27.4); 140 | 54.9 (27.6); 132 |
| Feels relatively wealthy compared to others (0-100, Strongly disagree) | 52.7 (24.5); 274 | 51.6 (24.1); 138 | 54.8 (24.1); 132 |
| Feels like they have enough money (0-100, Strongly disagree) | 54.0 (28.7); 273 | 55.1 (28.0); 137 | 55.5 (28.5); 132 |
| **Financial Strain* - n (%)** | **n=290** | **n=141** | **n=141** |
| Living comfortably | 113 (39.0) | 33 (23.4) | 33 (23.4) |
| Doing alright | 129 (44.5) | 52 (36.9) | 54 (38.3) |
| Just about getting by | 36 (12.4) | 29 (20.6) | 33 (23.4) |
| Finding it quite difficult | 6 (2.1) | 16 (11.3) | 13 (9.2) |
| Finding it very difficult | 1 (0.3) | 9 (6.4) | 6 (4.3) |
| Prefer not to say | 5 (1.7) | 2 (1.4) | 2 (1.4) |
| **Household composition* - n (%)** | **n=295** | **n=146** | **n=142** |
| Lives alone | 28 (9.5) | 26 (17.8) | 14 (9.9) |
| Lives with partner | 237 (80.3) | 97 (66.4) | 113 (79.6) |
| Lives with child/children | 126 (42.7) | 50 (34.2) | 68 (47.9) |
| Lives with parents | 14 (4.7) | 11 (7.5) | 12 (8.5) |
| Lives with friends | 2 (0.7) | 4 (2.7) | - |
| Other | 8 (2.7) | 5 (3.4) | 2 (1.4) |
| Household size - mean (SD); n | 2.7 (1.1); 288 | 2.5 (1.2); 140 | 2.9 (1.3); 141 |
| **Highest educational qualification* - n (%)** | **n=266** | **n=127** | **n=129** |
| Degree level or above | 134 (50.4) | 62 (48.8) | 53 (41.1) |
| Another kind of qualification | 132 (49.6) | 65 (51.2) | 76 (58.9) |
| **Employment Status* - n (%)** | **n=286** | **n=141** | **n=141** |
| Paid job - Full time (30+ hours per week) | 165 (57.7) | 83 (58.9) | 86 (61.0) |
| Paid job - Part time (8-29 hours per week) | 17 (5.9) | 8 (5.7) | 8 (5.7) |
| Paid job - Part time (Under 8 hours per week) | 2 (0.7) | - | 1 (0.7) |
| Self-employed | 22 (7.7) | 8 (5.7) | 18 (12.8) |
| Full time student | 3 (1.0) | 2 (1.4) | - |
| Unemployed and seeking work | 2 (0.7) | 5 (3.5) | 2 (1.4) |
| Retired | 66 (23.1) | 13 (9.2) | 19 (13.5) |
| Not in paid work due to illness or disability | 6 (2.1) | 18 (12.8) | 5 (3.5) |

| | No Mental Health Condition n=295 | Mental Health Condition n=146 | Latent Mental Health Condition n=142 |
|---|---|---|---|
| Not in paid work for other reason | 1 (0.3) | - | - |
| Other | 1 (0.3) | 4 (2.8) | 2 (1.4) |
| Prefer not to say | 1 (0.3) | - | - |
| **Access to self-monitoring equipment* - n (%)** | **n=293** | **n=144** | **n=142** |
| Owns scales for self-weighing | 251 (85.7) | 123 (85.4) | 113 (79.6) |
| Scales link to internet/app | 33 (12.9) | 20 (15.9) | 23 (19.3) |
| Owns an activity tracker/pedometer | 161 (54.9) | 78 (53.8) | 78 (54.9) |
| Highest weight (kg) - mean (SD); n | 120.6 (19.9); 286 | 125.7 (21.4); 141 | 131.6 (25.5); 139 |
| Lowest weight (kg) - mean (SD); n | 89.8 (15.7); 284 | 88.6 (16.9); 137 | 93.5 (20.8); 137 |
| Intended weight loss in study (kg) - mean (SD); n | 20.8 (14.9); 283 | 25.7 (16.6); 140 | 24.6 (13.8); 139 |
| Weight loss attempts - median (P25, P75); n | 5.0 (3.0-10.0); 286 | 7.0 (4.0-10.0); 141 | 10.0 (4.0-13.0); 140 |
| **Measured weight and height - n (%)** | **n=295** | **n=146** | **n=142** |
| Weight (kg) - mean (SD); n | 115.1 (18.0); 295 | 119.2 (20.4); 146 | 124.8 (21.6); 142 |
| Height (cm) - mean (SD); n | 176.5 (7.4); 295 | 176.9 (6.8); 146 | 178.7 (6.9); 142 |
| BMI (kg/m^2) - mean (SD); n | 36.9 (5.1); 295 | 38.1 (6.3); 146 | 39.1 (6.2); 142 |
| ≥30-<35; n (%) | 125 (42.4) | 55 (37.7) | 41 (28.9) |
| ≥35-<40; n (%) | 105 (35.6) | 48 (32.9) | 50 (35.2) |
| ≥40; n (%) | 65 (22.0) | 43 (29.5) | 51 (35.9) |

* Self-reported ^ Includes mental health condition

**S2 Table. Number (%) of mental health and wellbeing outcomes identifying a latent mental health condition in participants.**

| No. of outcome measures | Texts + Incentive (N=50) | Texts Alone (N=48) | Control (N=44) | Total (N=142) |
|---|---|---|---|---|
| 1 outcome measure | 28 (56.0) | 32 (66.7) | 30 (68.2) | 90 (63.4) |
| 2 outcome measures | 17 (34.0) | 15 (31.2) | 14 (31.8) | 46 (32.4) |
| 3 outcome measures | 5 (10.0) | 1 (2.1) | - | 6 (4.2) |

**S3 Table. Number of mental health and wellbeing baseline outcomes indicative of a latent mental health condition in participants (N, %) with a mental health condition by trial group.**

| No. of outcome measures | Texts + Incentive (N=51) | Texts Alone (N=46) | Control (N=49) | Total (N=146) |
|---|---|---|---|---|
| No outcome measure | 23 (45.1) | 17 (37.0) | 22 (44.9) | 62 (42.5) |
| 1 outcome measure | 13 (25.5) | 12 (26.1) | 15 (30.6) | 40 (27.4) |
| 2 outcome measures | 9 (17.6) | 3 (6.5) | 9 (18.4) | 21 (14.4) |

| | | | | |
|---|---|---|---|---|
| 3 outcome measures | 4 (7.8) | 9 (19.6) | 2 (4.1) | 15 (10.3) |
| 4 outcome measures | 2 (3.9) | 5 (10.9) | 1 (2.0) | 8 (5.5) |

**S4 Table. Number (%) of participants with a mental health condition (n=146) with mental health and wellbeing outcome measure scores indicative of a latent mental health condition by trial group.**

| N (%) | Texts + Incentive | Texts Alone | Control |
|---|---|---|---|
| WSSQ | 16 (11.0) | 19 (13.0) | 17 (11.6) |
| WEMWBS | 19 (13.0) | 22(15.1) | 9 (6.2) |
| PHQ-4 | 9 (6.2) | 15 (10.3) | 8 (5.5) |
| EQ-5D-5L-AD | 7 (4.8) | 9 (6.2) | 9 (6.2) |

* Rows are not mutually exclusive

### S5. Topic Guide

**12 Month Interview Topic Guide (SMS only and SMS+I Groups)**

**12M Questions and Prompts**

'I would like to have a chat with you about your experience of Game of Stones.'

**General**

- **How have you found taking part in Game of Stones?**
  (Probe further around their experience (usefulness) of the texts, the follow ups, the trial and incentives (incentive group)
  **Prompts:**
  - have there been any benefits for you from taking part in Game of Stones?
  - Any benefits that you didn't expect?
  - Have there been any downsides to it?
  - how did you feel about the weight-loss targets?
  - were the targets realistic for you? (Or probe around other words they use to describe targets)
  - if a friend was thinking of joining Game of Stones, what would you say to them?

- **Have you any suggestions about how Game of Stones, or parts of it, could be improved?**

***If not already covered:**

- **What about…the duration (texts stopping after 12M) …the texts …the website …incentives …getting to know other men?**

- **Has anything made it difficult for you to take part in Game of Stones?**

  *NB Examples of key words to probe further: stress, mental health, physical health, failure, shame, medicines, body image, self-esteem, relationships (e.g. you talked about Xxx can you tell me a bit more about that/ the impact that's had generally/ game of stones?)

  **Prompts:** pandemic (What kind of impact has Covid or the pandemic had on the Game of Stones for you?); cost of living crisis (What about the rises in costs that have happened recently – how have they impacted Game of Stones for you?)

**3 and 6 Month Follow-up Appointments**

- **Thinking back to your 3 and 6 months appointments – can you tell me about what they were like?**

**Prompts:**

- How were you on the run-up to these appointments?
- Did your behaviour or motivation change, before or after these appointments?
- How was the length of the appointment?

- **What did you think about the appointments being face-to-face?**
  **Prompts:**
  - Some men have had contact with the same researcher throughout Game of Stones and for others it's been different researchers – what was your experience?
  - Probe depending on experience (how did you feel about the fact it was the same person throughout?/ how did you feel about the fact it was different people?)
  - Would you change anything about these e.g. when they happened/ timeframe or who carried them out?

- **Some men found it difficult to attend appointments, did anything get in the way of you coming to both of your 3M and 6M appointments?**

**Prompts:**

- If yes, what would have helped you to attend?
- What might encourage other men to attend appointments?

- **In future, it might be difficult to have appointments for everyone, or at all times. Do you have any suggestions about what we might do instead?**

**Prompts:**

- How do you think that would work?
- What would the benefits/downsides be?
- What would you think about replacing these face-to-face appointments with digital scales?

**Potential impact on weight outcomes.**

- **Has anything happened in the last 12 months, outside of Game of Stones, that might have impacted your food or drink choices or physical activity?**

Prompts: Helpful or unhelpful experiences. If not already discussed:

- Have any health problems had an impact on your weight? (e.g. stress, mental health, physical health problems, family illness)
- Have your circumstances changed in any way that has impacted your weight? (e.g. impact of pandemic, cost of living, employment, relationships)

- **Is there anything that will help you to continue or support your efforts to manage your weight?**

**Prompts:** any strategies that you have found particularly useful? local weight management services, online programmes, apps

**Incentives (SMS+I group only)**

- **What did you think about being offered money for reaching weight loss targets?**

**Prompts:**

- How much money did you get? How did this affect your approach to losing weight? Did this change over time?
- During the study, the money for incentives was funded by the NIHR, the research arm of the NHS. Who else do you think might be able to provide it in future? Why do you say this?

**Final Questions:**

- **Game of Stones has been designed with men in mind how important was that for you?**

**Prompt:** Is there anyone else you think we should consider offering it to (e.g. some people have suggested it should continue as men only, be available to men with a health condition or on low incomes, be open to women)? Why do you say that?

- **Is there anything about your time on Game of Stones that you'd like to tell me about that I haven't asked?**

**Thank you for taking part!!**

**S6 Table. Description of change in mental health and wellbeing outcome measures between baseline and 12 months by mental health category.**

| | |
|---|---|
| **Men Living with a Mental Health Condition** | |
| **Wellbeing Measures** | WEMWBS scores increased between baseline and 12-months for the texts and incentive group (43.7 to44.63) and the texts alone group (40.37 to 43.18). WEMWBS scores decreased between baseline and 12-months for the control group (44.17 to 43.95). |
| | WSSQ scores increased between baseline and 12-months for the control group (37.54 to 39.52). WSSQ scores decreased between baseline and 12-months for the texts and incentive group (36.89 to 34.84) and for the texts alone group (38.5 to 36.7). |
| | EQ-5D-5L scores increased between baseline and 12-months for the texts and incentive (0.64 to 0.66), and control (0.64 to 0.71) groups. EQ-5D-5L scores decreased between baseline and 12-months for the texts alone (0.62 to 0.60) group. |
| | EQ-5D VAS scores increased between baseline and 12-months for the texts and incentive (57.20 to 67.67), texts alone (58.05 to 62.29) and control (59.77 to 62.10) groups. |
| **Screening Measures** | PHQ-4 scores decreased between baseline and 12-months for the texts and incentive (4.03 to 3.39), texts alone (4.46 to 4.03) and control (4.00 to 3.26) groups. |
| | EQ-5D-AD scores increased between baseline and 12-months for the texts alone (2.57 to 2.64) group. EQ-5D-AD scores decreased between baseline and 12-months for the texts and incentive (2.55 to 2.53) and control (2.69 to 2.33) groups. |
| **Men living with a Latent Mental Health Condition** | |
| **Wellbeing Measures** | WEMWBS scores increased between baseline and 12-months for the texts and incentive (40.82 to 45.84), texts alone (42.04 to 43.04) and control (40.28 to 44.27) groups. |
| | WSSQ scores decreased between baseline and 12-months for the texts and incentive (43.32 to 39.07), texts alone (40.95 to 37.90) and control (41.53 to 38.61) groups. |
| | EQ-5D-5L scores increased between baseline and 12-months for the texts and incentive (0.70 to 0.72), texts alone (0.70 to 0.71) and control (0.67 to 0.73) groups. |
| | EQ-5D VAS scores increased between baseline and 12-months for the texts and incentive (56.28 to 69.00), texts alone (57.68 to 65.65) and control (55.44 to 62.94) groups. |
| **Screening Measures** | PHQ-4 scores increased between baseline and 12-months for the control (2.78 to 2.88) group. PHQ-4 scores decreased between baseline and 12-months for the texts and incentive (3.22 to 2.63) and texts alone (3.86 to 3.04) groups. |
| | EQ-5D-AD scores increased between baseline and 12-months for the texts and incentive (2.02 to 2.03) group. EQ-5D-AD scores decreased between baseline and 12-months for the texts alone (2.19 to 2.07) and control (1.91 to 1.88) groups. |
| **Men without a Mental Health Condition or Latent Mental Health Condition** | |
| **Wellbeing Measures** | WEMWBS scores increased between baseline and 12-months for the texts and incentive (52.11 to 3.56) group. WEMWBS scores decreased |

| | |
|---|---|
| | between baseline and 12-months for the texts alone (52.10 to 51.06) and control (50.96 to 50.08) groups. |
| | WSSQ scores decreased between baseline and 12-months for the texts and incentive (31.02 to 30.62), texts alone (30.19 to 29.27) and control (31.81 to 30.94) groups. |
| | EQ-5D-5L scores increased between baseline and 12-months for the texts and incentive (0.79 to 0.84) and control (0.78 to 0.82) groups. EQ-5D-5L scores decreased between baseline and 12-months for the texts alone (0.82 to 0.78) group. |
| | EQ-5D VAS scores increased between baseline and 12-months for the texts and incentive (66.67 to 71.40), texts alone (67.59 to 73.35) and control (66.33 to 68.87) groups. |
| **Screening Measures** | PHQ-4 scores increased between baseline and 12-months for the texts and incentive (1.01 to 1.32), texts alone (0.90 to 1.55) and control (1.05 to 1.36) groups. |
| | EQ-5D-AD scores increased between baseline and 12-months for the texts alone (1.27 to 1.34) and control (1.50 to 1.57) groups. EQ-5D-AD scores decreased between baseline and 12-months texts and incentive (1.37 to 1.36) group. |
